# Emergence of a Novel Reassortant Clade 2.3.2.1c Avian Influenza A/H5N1 Virus Associated with Human Cases in Cambodia

**DOI:** 10.1101/2024.11.04.24313747

**Authors:** Jurre Y. Siegers, Ruopeng Xie, Alexander M.P. Byrne, Kimberly M. Edwards, Shu Hu, Sokhoun Yann, Sarath Sin, Songha Tok, Kimlay Chea, Sreyviseth Horm, Chenthearath Rith, Seangmai Keo, Leakhena Pum, Veasna Duong, Heidi Auerswald, Yisuong Phou, Sonita Kol, Andre Spiegel, Ruth Harvey, Sothyra Tum, San Sorn, Bunnary Seng, Yi Sengdoeurn, Chau Darapheak, Chin Savuth, Makara Hak, Vanra Ieng, Sarika Patel, Peter Thielen, Filip F. Claes, Nicola S. Lewis, Ly Sovann, Vijaykrishna Dhanasekaran, Erik A. Karlsson

**Affiliations:** Virology Unit, Institut Pasteur du Cambodge, Phnom Penh, Cambodia; School of Public Health, LKS Faculty of Medicine, The University of Hong Kong, Hong Kong SAR, China; HKU-Pasteur Research Pole, LKS Faculty of Medicine, The University of Hong Kong, Hong Kong SAR, China; Direction, Institut Pasteur du Cambodge, Phnom Penh, Cambodia; Worldwide Influenza Centre, The Francis Crick Institute, London, UK; National Animal Health and Production Research Institute, Phnom Penh, Cambodia; Communicable Disease Control Department, Ministry of Health, Phnom Penh, Cambodia; National Institute of Public Health, Ministry of Health, Phnom Penh, Cambodia; Food and Agriculture Organization of the United Nations (FAO) Country Office, Phnom Penh, Cambodia; World Health Organization Country Office, Phnom Penh; Johns Hopkins University Applied Physics Laboratory, Laurel, MD, USA; FAO Emergency Centre for Transboundary Animal Diseases, Regional Office for Asia and the Pacific, Bangkok, Thailand

**Keywords:** avian influenza, H5N1, Cambodia, Southeast Asia, One Health, genomic surveillance

## Abstract

After nearly a decade without reported human A/H5N1 infections, Cambodia faced a sudden resurgence with 16 cases between February 2023 and August 2024, all caused by A/H5 clade 2.3.2.1c viruses. Fourteen cases involved a novel reassortant A/H5N1 virus with gene segments from both clade 2.3.2.1c and clade 2.3.4.4b viruses. The emergence of this novel genotype underscores the persistent and ongoing threat of avian influenza in Southeast Asia. This study details the timeline and genomic epidemiology of these infections and related poultry outbreaks in Cambodia.

## II. Introduction

Avian influenza viruses (AIVs), particularly highly pathogenic avian influenza (HPAI) A/H5N1 strains, have caused significant economic losses in the poultry industry and pose a serious zoonotic threat (1). Monitoring AIV circulation and evolution in high-risk regions, such as the Greater Mekong Subregion (GMS), is crucial due to the region’s heavy reliance on agriculture, particularly backyard poultry farming, which often operates with minimal biosafety or biosecurity measures.

Cambodia, a lower middle-income country in the GMS has a large socio-economic dependence on agriculture and faces ongoing circulation of AIVs in poultry. Longitudinal surveillance at key live bird markets (LBMs) in Cambodia has shown year-round co-circulation of AIVs of agricultural and zoonotic concern including HPAI A/H5N1, low pathogenic avian influenza (LPAI) A/H9N2, and to a lesser extent, A/H7 viruses^1–4^. In addition, there has been infrequent detection of wild-bird-associated LPAI subtypes, including A/H1, A/H2, A/H3, A/H4, A/H6, A/H10, A/H11, and even A/H14, the latter marking the first detection of this subtype in Asia^5–7^. Along with the high prevalence of AIV in poultry, Cambodia has faced numerous instances of zoonotic spillover between 2005 and 2014, resulting in 21 human cases of A/H5N1, with 19 deaths (case-fatality rate [CFR]: 90.5%) by the end of 2012. In January 2013, a clade 1.1.2 reassortant virus emerged in poultry in the GMS, containing the hemagglutinin (HA) and neuraminidase (NA) genes from a clade 1.1.2 genotype Z virus and internal genes from a clade 2.3.2.1a virus. Following the emergence of this novel reassortant, 26 new human cases were reported in 2013 alone, with 15 deaths (CFR: 57.9%)^5,8^. The outbreak continued into the first quarter of 2014, resulting in 8 confirmed cases and 4 deaths (CFR: 50%)^9^. This clade 1.1.2 reassortant virus was replaced by clade 2.3.2.1c viruses in poultry by March 2014, with only one clade 2.3.2.1c human infection occurring thereafter^3^. The reassortant 1.1.2 virus has not been detected since. HPAI A/H5 clade 2.3.2.1c viruses have continued to circulate in the GMS since then^10^; however, clade 2.3.4.4b viruses were first detected in Cambodian LBMs in 2021, co-circulating with clade 2.3.2.1c viruses^11^.

After almost a decade without a human case, a cluster of A/H5N1 cases involving two related individuals was detected in February 2023 caused by the historically circulating clade 2.3.2.1c viruses (1 death, CFR: 50%). Between October 2023 and end of August 2024, Cambodia reported 14 more A/H5N1 cases (6 deaths, CFR: 42.8%). Critically, this recent surge in human A/H5N1 cases is linked to the emergence of a novel reassortant genotype, which combines elements from multiple AIV lineages. This study aims to elucidate the timeline, transmission dynamics, and evolutionary patterns of these A/H5N1 viruses across human and avian populations in Cambodia between February 2023 and August 2024. Here, we seek to trace viral origins, identify transmission pathways, and assess potential for increased risks to human health.

## III. Materials and Methods

### Ethical approval

All H5 sequences and information (e.g. collection date, location) included in this analysis were obtained as part of human H5 case testing and One Health response as described below. All data included in this study is available in the public record. All patient samples were de-identified and no other individual-specific information was used in this study. Analyses in this study have been approved by the Cambodian National Ethics Committee for Health Research (#365NECHR/2024).

### Sample collection

#### Human case detection

The Cambodian Influenza-like Illness (ILI) and Severe Acute Respiratory infections (SARI) surveillance systems are part of the WHO Global Influenza Surveillance and Response System and are a public health activity managed by the Ministry of Health (MoH) and Cambodia’s Communicable Disease Control (CCDC) Department. This surveillance system has existed since 2006, with increased capacity during the COVID-19 pandemic. Cases were also detected through event-based surveillance and active case finding.

#### Case investigation

For each human case, a collaborative One Health investigation was conducted by MoH-CCDC, the National Animal Health and Production Research Institute (NAHPRI), Ministry of Environment, and provincial authorities, with assistance from the United States Centers for Disease Control and Prevention (US CDC), the World Health Organization (WHO), and the Food and Agriculture Organization of the United Nations (FAO).

#### Animal sampling

Animal sampling was conducted by NAHPRI under the direction of the General Directorate for Animal Health and Production (GDAHP), Cambodian Ministry of Agriculture, Forestry and Fisheries (MAFF) along with provincial officials as part of One Health investigations surrounding each human case. In addition, Institut Pasteur du Cambodge (IPC) and NAHPRI/GDAHP collaborate on active disease surveillance activities focusing on LBMs^11^, with assistance from the Food and Agriculture Organization of the United Nations (FAO).

### RNA extraction and qRT-PCR

Viral RNA was extracted from samples using the QIAamp Viral RNA Mini Kit (Qiagen, Maryland, USA) according to the manufacturer’s protocol. Quantitative real-time reverse transcription PCR (qRT-PCR) was performed to screen for the matrix (MP) gene of influenza A virus, as described previously^6,7^. Samples with a cycle threshold (Ct) value <40 were deemed positive. Both original samples and isolates were subtyped using influenza RT-PCR assays to test for H1, H3, H5, H7 and H9 HA genes and N1, N2, N6, N8 and N9 NA genes^6,7^.

### Virus isolation

Virus isolation was attempted for all qRT-PCR positive human and poultry samples from each outbreak cluster^12^. Ten-day-old embryonated hens’ eggs were inoculated via the allantoic route and allantoic fluid was harvested three days post inoculation. For each sample, three blind passages were performed. Additionally, virus isolation from human samples was performed using Madin-Darby canine kidney (MDCK) cells stably transfected with human CMP-*N*-acetylneuraminate:β-galactoside α-2,6-sialyltransferase (SIAT1), which leads to overexpression of the α-2,6-linked sialic acid (α2,6-SA) receptor^13^. The presence of influenza virus in the allantoic fluid and cell culture supernatant was tested using qRT-PCR as described above.

### Sequencing

Whole viral genomes were amplified from patient nasal swabs, poultry samples, and/or isolates using a modified multi-segment RT-PCR method that includes integrated molecular indices (iiMS-PCR)^14^. The iiMS-PCR products were then pooled, prepared for sequencing with the ligation sequencing kit SQK-LSK114 (R10 chemistry, Oxford Nanopore Technologies (ONT), Oxford, UK) and sequenced on the GridION platform (ONT) using R10 flow cells as described previously^15^. Sequencing reads were de-multiplexed, quality trimmed, and filtered using Porechop software (https://github.com/rrwick/Porechop). Consensus sequences were generated using CDC’s Iterative Refinement Meta Assembler (IRMA) v1.1.4 using the default “IRMA FLU-minion” settings^16^. Consensus sequences were manually inspected for errors such as insert-deletion mutations (INDELs) and mixed bases and corrected if required. A minimum coverage depth of 10x was set for all genes. Genomic sequencing was completed for twelve of the fifteen reported human cases, with one HA sequence (A/Cambodia/RL240007/2024) being partial. This study generated a total of 83 gene segments obtained from 12 human A/H5N1 and 329 segments from 51 avian A/H5N1 influenza A viruses were deposited in GISAID. Accession numbers can be found in supplemental table 2.

### Phylogenetic analysis

All available full-genome H5Nx influenza A virus sequences (N = 11,765), including the internal protein coding genes were obtained from GISAID EpiFlu^17^ (26) databases (accessed on 2024-03-21). The full genome sequences were concatenated using SeqKit^18^ and aligned prior to the inference of preliminary maximum-likelihood (ML) phylogenetic trees with Augur^19^. This tree was then down-sampled to maintain phylogenetic representatives using PARNAS^20^. Sequences generated in this study, along with publicly available sequences from Asia since 2020-01-01, were added to this dataset to enhance regional representation and maximize phylogenetic diversity. Sequences identified as duplicates (based on strain name), laboratory-derived, mixed subtypes, or with coverage <90% of full length were excluded from further analysis. Each gene dataset was aligned with MAFFT v.7.490^21^, and optimized manually. Large-scale ML phylogenetic trees were inferred with FastTree v2^22^ using the GTR nucleotide substitution model. Focused ML trees were generated in IQ-TREE v.2.2.0^23^ using the best-fit nucleotide substitution model (HKY_+_F_+_G4) chosen according to Bayesian Information Criterion. Trees were visualized and annotated using FigTree v.1.4.4 (http://tree.bio.ed.ac.uk/software/figtree/) and ggTree^24^ in R (version 4.2.2).

### Reassortment analysis

To investigate reassortment events among major clades (2.3.2.1c, 2.3.4.4b, 2.3.4.4x, 2.3.2.1a, 2.3.4, 1.1.2) and minor clades since 1996, we first constructed a ML tree for HA gene sequences using IQ-TREE v.2.1.4. We subsampled 1,000 genomes, ensuring all Cambodian sequences were retained, using the Phylogenetic Diversity Analyzer tool v.1.0.3 (http://www.cibiv.at/software/pda). Subsequently, we constructed ML trees for the remaining seven segments, including polymerase basic 2 (PB2), polymerase basic 1 (PB1), polymerase acidic (PA), neuraminidase (NA), nucleoprotein (NP), matrix (MP), and non-structural (NS) gene segments of the 1,000 subsampled genomes. Baltic v.0.1.5 (https://github.com/evogytis/baltic) was used to visualize the incongruence between the phylogenetic trees across the eight genes. The HA tree was rooted using clade 0, while the other trees were rooted at the midpoint.

### Molecular marker analysis

Molecular markers were analyzed using a pipeline described previously^25^ and Flutile (https://github.com/flu-crew/flutile).

## IV. Results

### Direct poultry-to-human transmission with no evidence of human-to-human spread

Viruses sequenced from human cases in February 2023 in Prey Veng province were closely related to A/H5N1 viruses from poultry in Cambodian LBMs between 2022 and 2023, particularly from Phnom Penh and Kandal provinces (Figure 1a-c). Similarly, viruses from human cases in October 2023 (Svay Rieng and Prey Veng province), November 2023 (Kampot province), January 2024 (Siem Reap province), February 2024 (Kratie province), and early July 2024 (Svay Rieng province) were genetically similar to poultry viruses collected at their respective outbreak sites. Although the January 2024 Prey Veng human case lacked direct poultry-associated sequences, it was related to viruses detected from LBMs in Prey Veng and Kandal in January 2024. In February 2024, no viral sequences could be obtained from neither the Kampot human case nor poultry associated cases. In July 2024, no human viruses could be sequenced from the two Takeo province cases, although the two poultry-associated viruses were closely related to human and poultry viruses from the October 2023 Svay Rieng and one duck virus from the October 2023 Prey Veng outbreak. Investigations into the August 2024 cases Prey Vengcase showed that this virus was related to human and poultry viruses from October 2023 (Prey Veng and Svay Rieng) and July 2024 (Kampot). These findings consistently suggest zoonotic transmission from poultry to humans, with no evidence of human-to-human transmission (Figure 1c).

**Figure 1.**
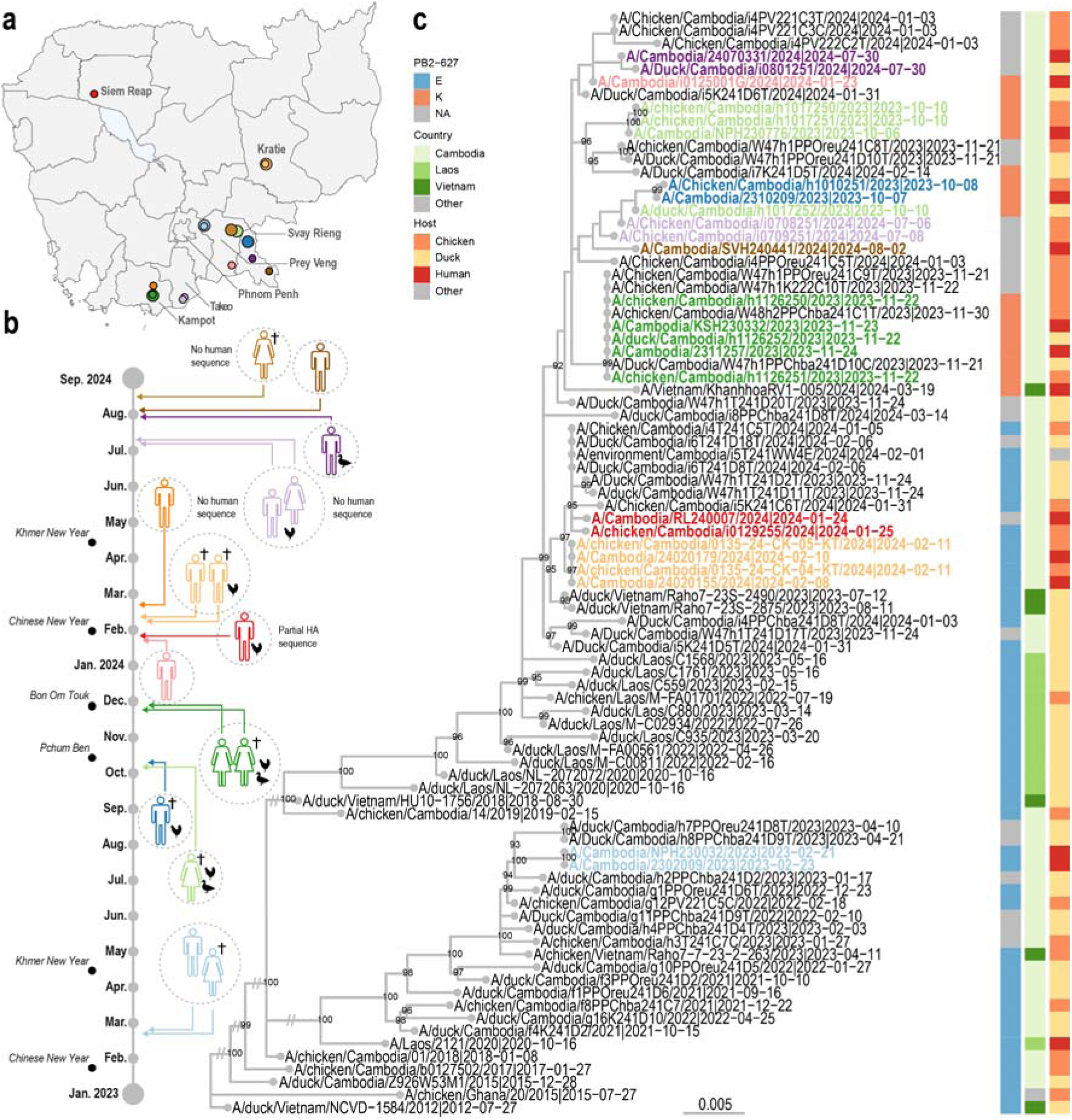
Geographic location, timeline of events, and phylogenetic relationships of human A/H5N1 infections and poultry-associated clade 2.3.2.1c A/H5N1 viruses in Cambodia, February 2023 - July 2024. a) Map of human A/H5N1 cases. Circles indicate clinical outcome, with smaller circles for recovered individuals and larger circles for fatal cases, colored by outbreak cluster. b) Timeline of A/H5N1 human cases. Colors represent outbreak clusters; dotted circles indicate epidemiologically linked cases; mortality symbols indicate fatal cases; chicken and duck silhouettes denote associated poultry viruses; and major Cambodian festivals are labeled. c) Maximum likelihood phylogenetic tree of the A/H5 HA genes, with human and poultry-associated A/H5N1 viruses colored by outbreak cluster. The color-coded right y--axes represent the E/K amino acid in the PB2 gene at position 627 (blue/orange), country of isolation (shades of green) and host (shades of red), respectively. Bootstrap values >80% are indicated. Scale bar denotes nucleotide substitutions per site. Tree is rooted to the clade 2.3.2.1c vaccine strain, A/duck/Vietnam/NCVD-1584/2012. Names are provided as strain_name | collection_date.

### Genotype replacement by a novel reassortant clade 2.3.2.1c virus in late 2023

Phylogenetic analysis of the HA gene segment reveals that all A/H5 viruses sequenced from human cases belong to clade 2.3.2.1c (Figure 1c). The virus responsible for the initial cases in February 2023 was closely related to clade 2.3.2.1c A/H5N1 viruses circulating in Cambodian poultry and wild birds since 2013^1^. These early viruses retained complete gene cassettes within clade 2.3.2.1c viruses (Figure 2). However, starting in late 2023 (October), HA sequences from both humans and poultry in Cambodia displayed significant divergence from earlier Cambodia strains. These HA genes clustered with two duck samples from Vietnam during July and August 2023, and were derived from a sublineage of clade 2.3.2.1.c continuously detected in Laos during October 2020 and May 2023. Together, this indicates an introduction of a sublineage of clade 2.3.2.1c viruses with a common ancestor detected in poultry in Vietnam and Laos, but the exact introduction pathway cannot be pinpointed due to gaps in available surveillance data.

**Figure 2.**
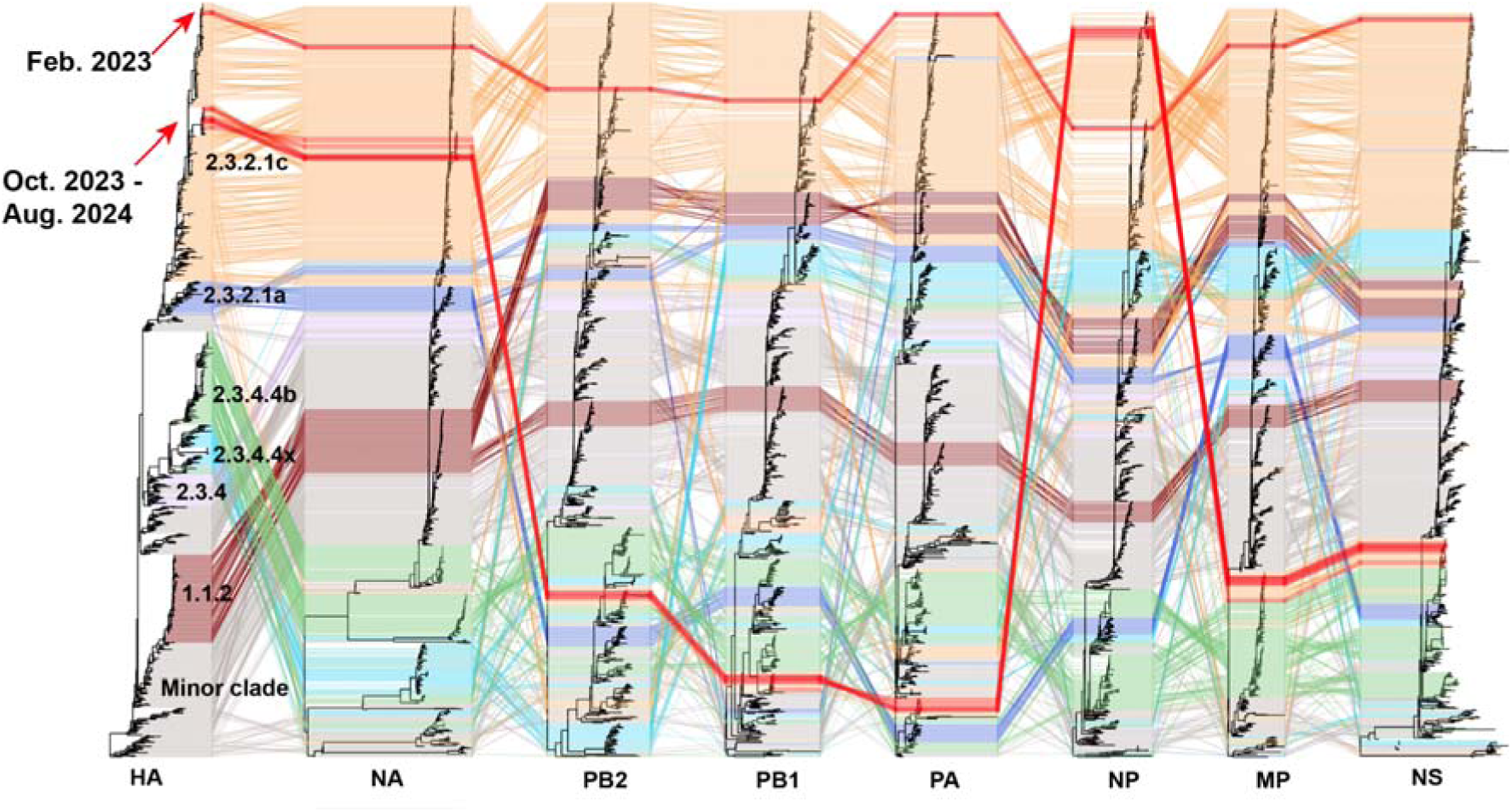
Genotypic reassortment patterns of clade 2.3.2.1c A/H5N1 viruses causing human infections in Cambodia since February 2023. Tanglegram of ML phylogenetic trees for each genomic segment. For each virus, links have been displayed to connect phylogenetic positioning of individual viruses across all eight genomic segments. Phylogenetic links are color coded based on relevant A/H5 clade numbering: clade 2.3.2.1c in orange, 2.3.2.1a in dark blue, 2.3.4.4 in light blue, 2.3.4.4b in green, 2.3.4 in purple, 1.1.2 in brown, minor A/H5N1 clades in grey and recent Cambodian human A/H5N1 sequences in bright red.

Phylogenetic analyses of each of the gene segments revealed that A/H5N1 viruses in Cambodia, detected from October 2023 onwards in both humans and poultry, represent a novel genotype resulting from reassortment. This reassortment combines segments from clade 2.3.2.1c (HA, NP, and NA) and clade 2.3.4.4b (PB2, PB1, PA, MP, and NS) viruses (Figure 2). In Cambodia, this novel 2.3.2.1c genotype has completely replaced the endemic 2.3.2.1c genotype that has dominated in poultry for the last decade.

### Molecular markers of risk in novel reassortant genotype Cambodian A/H5N1 viruses have the potential for increased mammalian adaptation

Gene segments from the Cambodian human A/H5N1 viruses were analyzed for adaptive mutations affecting receptor binding affinity, HA fusion and/or stability, polymerase activity, virus replication in mammalian cells, and pathogenicity (Supplemental Table 1) ^26^. All human A/H5N1 viruses isolated since 2023 are classified asHPAI due to the presence of a multi-basic cleavage site in the HA gene^27^. Three different cleavage sites were present: PQKERRKR↓GLF in human cases in February 2023, PQRERRKR↓GLF in cases from October 2024 to January 2024 and from July 2024 onwards, and KERRRKKR↓GLF, containing two additional basic amino acids from cases in February 2024.

Among the minimal set of five substitutions that confer airborne transmissibility in ferrets for the HPAI A/H5N1 virus strain A/Indonesia/5/2005 (E627K in PB2; H99Y in PB1; H103Y, T156A, and either Q222L or G224S in HA)^28,29^, only HA 156A was consistently observed in all A/H5N1 viruses from Cambodian human cases. Notably, a subset of the novel genotype forms a monophyletic lineage in the HA tree (arrow in Fig 1c, bootstrap value 92), and contains PB2 E627K, a key molecular determinant for host range, cross-species transmission and airborne transmission (PMID: 8445709 and 22723413)None of the human viruses from Cambodia contained previously defined HA substitutions (Q222L and G224S (H5 HA numbering) that switch receptor specificity from avian α2,3-SA to human α2,6-SA^30^. The presence of PB2 E627K and HA T156A substitutions, along with the potential for other functionally similar mutations, underscores the need for close monitoring of these A/H5N1 strains for signs of potential increased mammalian adaptation^31^.

## V. Discussion

Reassortment events leading to the exchange of gene segments between viruses can significantly alter viral properties and potentially increase the risk for spillover and human infections^32–35^. AIVs with novel gene combinations have been associated with increases in zoonotic human cases, such as with A/H5N1 (Cambodia clade 1.1) A/H7N9^34^, A/H3N8^33^, and even human pandemics^32^. Clade 2.3.4.4b A/H5N1, which has spread globally since its emergence in 2020, has undergone further reassortment during its dissemination.

The resurgence of zoonotic A/H5N1 cases in Cambodia underscores the complex dynamics of AIV evolution and potential for zoonotic transmission. Phylogenetic analysis identified two distinct spillover phases. The first phase, in February 2023, involved spillover of a clade 2.3.2.1c virus genotype, which had been dominant in Cambodian poultry since 2014^3^. The second phase, starting in October 2023, is marked by a novel reassortant virus combining genes from clades 2.3.2.1c and 2.3.4.4b. The exact origins of this reassortment are unclear, but likely facilitated by high-density poultry farming, wild bird migration, and cross-border poultry trade in the region, highlighting the ongoing risk of zoonotic AIV transmission in Southeast Asia.

While the phenotypic contributions of newly introduced clade 2.3.4.4b internal gene segments have yet to be elucidated, the presence of amino acid mutations in both human and poultry viruses such as E627K in the 2.3.4.4b-origin PB2 gene segment suggestsenhanced capacity for mammalian infection. To better understand the zoonotic risk that these viruses pose, further risk assessment *in silico*, *ex vivo*, *in vivo*, and *in vitro* is critical. In addition, the detection of the PB2 627K mutation in poultry is also a concern, as it may become established in widespread circulation.

The success of the Cambodian One Health response to these recent reassortant A/H5N1 cases highlights the importance of coordinated efforts between human and animal health sectors in managing zoonotic threats. In addition, rapid genomic surveillance has been critical in understanding the dynamics of virus spillover events and has provided clear evidence linking human cases to infected poultry from the same household. This integration of genomic data with field investigations allowed for timely identification of transmission pathways and underscores the importance of ongoing surveillance in high-risk areas. These efforts exemplify the strength of the One Health approach in Cambodia, where collaboration across disciplines and rapid data sharing have proven essential for understanding, responding and controlling human AIV outbreaks in the country.

Given the success of Cambodian One Health response efforts being able to link cases to direct contact with infected poultry, public health strategies should prioritize reducing human exposure, particularly in high-risk rural areas. Reinforcing public education on the risks associated with direct contact with infected birds has proven effective during other AIV outbreaks, but significant challenges remain in achieving consistent behavioral changes. Most Cambodians in rural areas are still at high-risk for potential exposure to AIV^36^. In conjunction with human case investigations, community-based education campaigns have been implemented in Cambodia, but their reach and effectiveness vary^37,38^. Only 32% of survey participants reported receiving information about AIV from healthcare providers, 10.6% from village health support groups, and 2% from village animal health workers (VAHW)^37^. Furthermore, only 49% of participants reported poultry illness and deaths to local authorities, and 23% of participants reported that they cook sick or dead poultry for consumption. This is particularly crucial given that 68.3% of participants raised chickens in their backyards and 10.2% raised ducks, generating a high level of potential exposure to infected birds^37^. Further research is necessary to identify, understand and overcome the barriers in risk perception that hinder individuals from adopting safer behaviors, and to more effectively tailor behavioral messages to specific target audiences. Educational interventions on the necessity of early access to healthcare, recognizing AIV infection as a life-threatening disease, and awareness of the risks associated with handling sick poultry have proven to be a powerful tool in increasing awareness and modifying health behaviors^40,41^.

Training and funding for VAHW and primary healthcare workers (PHW) are critical for early detection and outbreak response^40–44^. Educational interventions can help VAHW and PHW identify poultry deaths and exposure history, while also supporting event-based surveillance in both human and animal sectors. Strengthening VAHW reporting and raising clinical suspicion among healthcare providers will enhance early detection and case management. Equipping healthcare workers to manage cases effectively and increasing awareness among high-risk individuals about early healthcare access and poultry-related risks are essential to improving preparedness^40,41^.

This study is limited by potential biases in human sample collection due to potential underreporting of cases from rural areas with limited healthcare access. Additionally, sustainable funding for active AIV surveillance is lacking across the GMS and broader Asia, resulting in inconsistent data collection and limited genomic information available for in-depth analysis. Moreover, there is an urgent need to develop a classification system that better reflects genotypes, as the conventional HA and NA subtype nomenclature fails to capture the full complexity of reassortant AIV genomes and associated evolutionary context.

In conclusion, these recurrent zoonotic infections caused by a novel reassortant A/H5N1 viruses in Cambodia serve as a reminder of the ever-present threat of AIV to global health security. Despite the recent focus on global dissemination and expanded host range of clade 2.3.4.4b ^45–47^, clade 2.3.2.1c viruses remain a significant concern, particularly in Asia, where the two clades co-circulate. A coordinated, regional approach is essential for effectively monitoring the threat of AIV and ensuring preparedness and response to emerging viral threats. Indeed, this study underscores the critical need and effectiveness of a unified, One Health approach to combat the evolving landscape of AIV. The urgency of this collaborative effort cannot be overstated as these viruses continue to adapt and reassort, increasing the risk of a strain evolving the capacity for efficient human-to-human transmission. To stay ahead of this threat, we must prioritize sustainable funding for long-term surveillance, enhance laboratory capacity for rapid whole genome sequencing, and foster open, trust-based information sharing across borders. Our collective preparedness today will determine our ability to protect global health tomorrow.

## Data Availability

This study generated a total of 83 gene segments obtained from 12 human A/H5N1 and 329 segments from 51 avian A/H5N1 influenza A viruses were deposited in GISAID. Accession numbers can be found in Supplemental Table 2.

## Acknowledgments

The investigators thank everyone involved in the critical discussions and review of this manuscript. They also thank everyone involved in influenza surveillance and response in the Kingdom of Cambodia including teams at the National Institute for Public Health, Cambodian Communicable Disease Control Department, Ministry of Health, National Animal Health and Production Institute, General Directorate of Animal Health and Production, Ministry of Agriculture, Forestry, and Fisheries, World Health Organization, provincial health directors, province rapid response teams, and the Virology Unit at Institut Pasteur du Cambodge who contributed to this study. We gratefully acknowledge the authors from the originating laboratories responsible for obtaining the specimens and the submitting laboratories where genetic sequence data were generated and shared via the GISAID Initiative, on which this research is based (full list of GISAID acknowledgments are available in supplemental material). We also thank all National Influenza Centers and laboratories that have supplied influenza viruses to the WHO Collaborating Centers for Reference and Research on Influenza for analysis. The text as published does not necessarily represent the official view of WHO or FAO.

## VI.#Funding Statement

Avian influenza work in the Virology Unit at Institut Pasteur du Cambodge was funded, in part, by the Food and Agriculture Organization of the United Nations, the World Health Organization, and the Bill and Melinda Gates Foundation. H.A. is supported, in part, by the German Centre for International Migration and Development. P.M.T was supported by Johns Hopkins APL internal research and development. R.X., K.M.E., S.H. and V.D. were funded in part by the National Institute of Allergy and Infectious Diseases, National Institutes of Health, United States Department of Health and Human Services, under Contract No. 75N93021C00016. The funders had no role in study design, data collection and interpretation, or the decision to submit the work for publication.

## VII.#Author Contributions

Conceptualization: J.Y.S., R.X., A.M.P.B., K.M.E., N.S.L., V. Dhanasekaran, and E.A.K. Data curation: J.Y.S., R.X., A.M.P.B., K.M.E., S.H., S.Y., S. Sin, S. Tok, K.C., S.V.H., C.R., S. Keo, L.P., H.A., Y.P., S. Kol, R.H., C.D., C.S., V.I., S.P., V. Dhanasekaran, and E.A.K. Formal analysis: J.Y.S., R.X., A.M.P.B., K.M.E., S.H., S.Y., S. Sin, S. Tok, K.C., S.V.H., C.R., S. Keo, L.P., H.A., Y.P., S. Kol, R.H., N.S.L., V. Dhanasekaran, and E.A.K. Funding acquisition: V. Duong, A.S., V.I., S.P., F.F.C., N.S.L., V. Dhanasekaran, and E.A.K. Investigation: J.Y.S., R.X., A.M.P.B., K.M.E., S.H., S.Y., S. Sin, S. Tok, K.C., S.V.H., C.R., S. Keo, L.P., Y.P., S. Kol, R.H., S. Tum, S. Sorn, B. Seng, Y.S., C.D., C.S., M.H., V.I., S.P., P.T., F.F.C., N.S.L., L.S., V. Dhanasekaran, and E.A.K. Methodology: J.Y.S., R.X., A.M.P.B., K.M.E., H.A., P.T., N.S.L., V. Dhanasekaran, and E.A.K. Project administration: V. Duong, H.A., A.S., S. Tum, S. Sorn, B. Seng, C.D., C.S., M.H., V.I., S.P., F.F.C., N.S.L., L.S., V. Dhanasekaran, and E.A.K. Resources: V. Duong, H.A., S. Tum, S. Sorn, B. Seng, C.D., C.S., M.H., V.I., S.P., P.T., F.F.C., N.S.L., L.S., V. Dhanasekaran, and E.A.K. Software: P.T. and V. Dhanasekaran Supervision: V. Duong, H.A., A.S., S. Tum, S. Sorn, B. Seng, Y.S., C.D., C.S., M.H., V.I., S.P., F.F.C., N.S.L., L.S., V. Dhanasekaran, and E.A.K. Validation: J.Y.S., R.X., A.M.P.B., K.M.E., S.Y., S.V.H., V. Dhanasekaran, and E.A.K. Visualization: J.Y.S., R.X., A.M.P.B., K.M.E., V. Dhanasekaran, and E.A.K. Writing - original draft: J.Y.S., R.X., A.M.P.B., K.M.E., P.T., F.F.C., N.S.L., V. Dhanasekaran, and E.A.K. Writing - review & editing: J.Y.S., R.X., A.M.P.B., K.M.E., S.H., S.Y., S. Sin, S. Tok, K.C., S.V.H., C.R., S. Keo, L.P., V. Duong, H.A., Y.P., S. Kol, A.S., R.H., S. Tum, S. Sorn, B. Seng, Y.S., C.D., C.S., M.H., V.I., S.P., P.T., F.F.C., N.S.L., L.S., V. Dhanasekaran, and E.A.K.

## VIII.#Conflict Of Interest

The authors declare no conflicts of interest.

## IX. Supplemental Figures and Tables

**Supplemental Table 1.** Molecular marker analysis and amino acid variation between human A/H5N1 virus in Cambodia, 2023-2024.

**Supplemental Table 2.** GISAID accession numbers.

